# Study on the Mechanism of Age Discrimination Perception on Self-Rated Health of the Elderly: Mediating Effect Based on Aging Attitude and Moderating Effect of Community Welfare

**DOI:** 10.1101/2025.07.09.25331167

**Authors:** Zhong Yuying, Xu Xianchi

## Abstract

**Background:** In practice, the construction of the old-age security system in China is not sufficient, and the mental health protection for the elderly is still lacking, which cannot effectively solve the health losses suffered by the elderly due to age discrimination. Although age discrimination is an external evaluation of the elderly, this evaluation will also potentially affect the elderly’s views on themselves and others’ aging, and then affect their health. In addition, building an elderly-friendly community is a trend in many places in recent years, which is expected to play an important regulatory role in the process of age discrimination perception affecting the health of the elderly.

**Objective:** This paper focuses on the influence of age discrimination perception on the self-rated health of the elderly and the intermediary role of aging attitude in this process, as well as the regulatory role played by community welfare in the first half of the intermediary effect.

**Methods:** Structural equation modeling (SEM) was used to analyze data from the China Longitudinal Aging Social Survey (CLASS) 2020 (N=6,066). Latent variables for age discrimination perception were constructed, and moderated mediation analysis was performed.

**Results:** Age discrimination has a negative impact on the self-rated health of the elderly. The perception of age discrimination has a negative impact on the self-rated health of the elderly by affecting their aging attitude. Community welfare moderates the influence path of age discrimination perception on aging attitude. Specifically, higher community welfare buffers the negative impact of age discrimination perception on aging attitude.

**Conclusions:** The validated model effectively identifies the mechanism of age discrimination perception on elderly health. This tool can aid policymakers in developing personalized strategies for aging populations. Community welfare interventions are crucial for mitigating the adverse effects of age discrimination.

## Introduction

The widespread age discrimination seriously damages the physical and mental health of the elderly. There are various types of discrimination in society, including racial discrimination, gender discrimination, age discrimination, etc. (Nelson, 2005). The discussion on racial discrimination and gender discrimination has been extensive, while the attention on age discrimination is relatively less. Age discrimination is one of the most common forms of discrimination (Palmore, 1999), and almost one third of the elderly reported that they had experienced age discrimination (Rippon & Steptoe, 2015). A large number of studies have shown that the perception of age discrimination will have a negative impact on the physical and mental health of the elderly (Jackson S E et al., 2019; Levy B R et al., 2023).

Old-age security should meet the multi-level and multi-dimensional needs of the elderly, but in fact, China’s old-age security system ignores the spiritual security for the elderly (Ran Xiaoxing and Chen Gong, 2023), which cannot effectively solve the health losses suffered by the elderly because of age discrimination. The Fifth Plenary Session of the 19th Central Committee of the Communist Party of China (CPC) officially put "implementing the national strategy of actively responding to population aging" into the national policy discourse system. The "Tenth Five-Year Plan" for the National Development of the Cause for the Aged and the Plan for the Service System for the Aged, further establishes the "active view of the aged" as a basic concept that runs through economic and social development, and requires it to be integrated into the whole process of economic and social development. Breaking age discrimination has become the consensus of action in implementing positive coping strategies.

As the basic unit of society, community plays an important role in the daily life of the elderly. In today’s aging, the continuous growth of the elderly population will bring a series of challenges to the community. With the decline of basic activity ability and cognitive ability, the needs of the elderly for community services and environmental resources are becoming more and more diversified, especially the realization of the "in-situ pension" model, which requires the empowerment of the community. Community represents the nearest peripheral social forces, and community welfare is closely related to the health of the elderly. The health of the elderly can be effectively improved through the improvement of community services. However, at present, the construction and excavation of community welfare in China is not sufficient. Under the background of promoting the construction of elderly-friendly communities, community welfare is expected to play a regulatory role in the process of age discrimination affecting health.

Previous studies have made a meaningful discussion on the influence of age discrimination perception on the health of the elderly, but the excavation of its influence mechanism is not sufficient and systematic. It is not conducive to our understanding of the internal mechanism of age discrimination on the health of the elderly, and it is not conducive to our taking corresponding intervention measures. Based on this, this paper focuses on the influence of age discrimination perception on the self-rated health of the elderly and the intermediary role of aging attitude in this influence path, and further discusses the regulatory role played by community welfare factors in the first half of the intermediary effect.

## literature review

Ageism, as a multidimensional social phenomenon, was first proposed by Butler to refer to the biased cognition of one age group to another (Butler, 1969). The World Health Organization further defines it as self-discrimination or discrimination by others based on age differences. Age discrimination poses a serious threat to the physical and mental health of the elderly: when the elderly think that they are a social burden, they may reduce their sense of life value and increase their risk of depression. Empirical studies have revealed that the incidence of chronic pain in the elderly with negative age stereotypes is 32% higher than that in the group with positive attitude in the next seven years (Levy Brr et al., 2023); And the probability of self-rated health status of age discriminators is significantly improved (Jackson S E et al., 2019). The research on the effect path of age discrimination on health is relatively insufficient. The Global Report on Age Discrimination issued by WHO points out that age discrimination damages the health of the elderly by reducing the quality of life, intensifying social isolation, inhibiting self-expression and increasing the risk of violence exposure. Stokes and others suggested that the decline of happiness may be an important mechanism (Stokes J E & Moorman S M, 2020); Marquette et al. found that negative age stereotype and discrimination perception will weaken self-esteem through cognitive deterioration of aging (Marquet M et al., 2019), and then damage health. It can be seen that age discrimination will have a negative impact on the physical and mental health of the elderly by affecting the quality of life, psychological state and happiness.

Laidlaw defines aging attitude as an individual’s view of aging of himself and others, which is also called "aging self-cognition" and "aging self-stereotype" (Laidlaw, 2007). Levy pointed out that it originated from the stereotype theory (Levy, 2009), which holds that the aging attitude comes from the stereotype of the elderly, which gradually formed and internalized from infancy. The aging attitude is mainly about the views and experiences of the elderly about themselves or others getting old (Ji Jingyao et al., 2018). Aging attitude is an important variable affecting the health of the elderly. Studies have confirmed that elderly people with positive aging attitude show better health status (Wurm et al., 2007); On the contrary, negative aging attitude significantly increases the prevalence of chronic diseases, the risk of physical dysfunction and the incidence of diseases in the elderly (Jang et al., 2004;; Warmoth et al., 2018). Related studies have confirmed that aging attitude plays an intermediary role in the process of other factors affecting the health of the elderly. Recently, it has been found that aging attitude plays a significant mediating role in the influence path of intergenerational support on depressive symptoms of the elderly (Shi Jiaming and Liu Xiaoting, 2024). Another study found that aging attitude plays an intermediary role in the process of cognitive function and mental health of rural elderly (Yin Lanxin et al., 2023).

China began to implement community welfare in the 1980s, and many studies have paid attention to the variable of community welfare. In the related research on the health of the elderly, community welfare has important research significance (Lv Xuanru et al., 2022; Berke et al., 2007). Many elderly people have the will to provide for the aged in situ, and some of their daily needs can be met through the community. Community welfare plays an important role in the life of the elderly, and rich community welfare can also provide the elderly with rich life experiences in their later years. The old-age care service for the elderly, whether it is national policy or existing research, practical or theoretical, is actually a kind of community welfare promoted by community forces. Community represents the nearest peripheral social forces, and community welfare is closely related to the elderly. Through the improvement of community services, the health of the elderly can be effectively improved.

To sum up, at present, the research on the influence of age discrimination perception on the health of the elderly has been rich, but there is a lack of in-depth discussion on its influence mechanism. Aging attitude will have an important impact on the health of the elderly, but no research has paid attention to the intermediary role of aging attitude factors in the process of age discrimination perception and health of the elderly. In addition, the factors of community welfare are closely related to the health of the elderly. Under the background of advocating the construction of elderly-friendly communities, the excavation and construction of community welfare in China still needs to be deepened. This paper focuses on the influence mechanism of age discrimination perception on the health of the elderly. What factors will regulate it? On the basis of existing research, this paper focuses on the influence of age discrimination perception on the self-rated health of the elderly and the intermediary role of aging attitude; At the same time, the moderating effect of community welfare on the path of age discrimination perception affecting aging attitude is discussed.

## The research framework

### Ethics statement

#### 1. Social exclusion theory

The concept of "social exclusion" first appeared in the French academic circle Silver H in 1970s, 1994; Martin C, 1996), whose core concern is to reveal how the vulnerable groups are systematically marginalized in key areas such as labor market access and social security resource allocation, and eventually become isolated groups. There are different types and manifestations of social exclusion, and age discrimination is one of them. Social exclusion theory can well explain the formation of age discrimination and how the perception of age discrimination affects the aging attitude and physical and mental health of the elderly. The elderly are excluded from the labor market because of their age and declining physical function, and suffer from unequal treatment in social mechanism, social distribution and social relations, and are excluded from the mainstream society (Li Bin, 2002). This situation will reduce the sense of self-worth of the elderly, and then affect their views on how to grow old and how others will eventually affect their physical and mental health.

#### 2. Community governance theory

The theory of community governance was put forward under the framework of governance theory, which was put forward by Mr. Xia Jianzhong. He believed that community governance refers to that multiple community governance subjects in the community jointly solve social public problems in the community through cooperation and consultation, and meet the individualized and reasonable needs of community residents. The essence of community governance theory is to emphasize the participation of multi-agents in the construction and governance of communities, and to meet the welfare needs of community residents in terms of goods, services, environment and other software and hardware.

Based on social exclusion theory and community governance theory, this paper puts age discrimination perception, aging attitude and elderly health into the same analytical framework and establishes a theoretical analysis model. This paper constructs the theoretical framework shown in Figure 1.

**Fig. 1.**
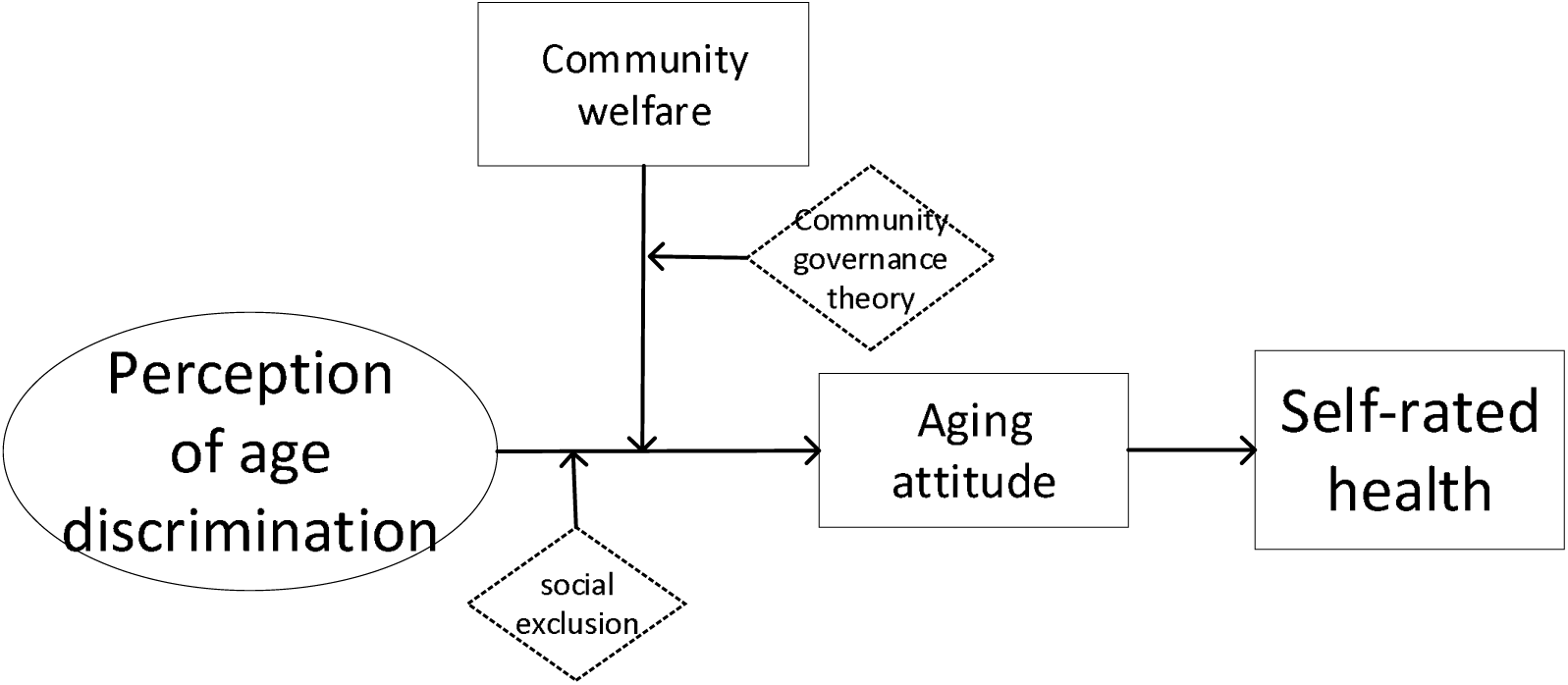
theoretical analysis frame diagram.

### Research hypotheses

According to the theory of social exclusion, the elderly are excluded from the labor market, treated unequally in terms of social mechanism, social distribution and social relations, and marginalized by the mainstream society, and become increasingly lonely and helpless groups (Li Bin, 2002). As a kind of social exclusion, age discrimination produces and strengthens the fear and slander of the aging process, as well as the stereotype of declining ability and being protected. Age discrimination seriously damages the physical and mental health of the elderly. When the elderly perceive themselves as a social burden, their sense of value will be seriously weakened, thus increasing the risk of depression. Studies have shown that the elderly with negative age stereotypes are much more likely to suffer from chronic pain than those with positive age stereotypes (Levy Brr et al., 2023); At the same time, patients who perceive age discrimination are more likely to report their general or poor health (Jackson S E et al., 2019). Accordingly, this study puts forward research hypothesis 1:

Research hypothesis 1: Age discrimination perception will negatively affect the self-rated health of the elderly. The more serious the perception of age discrimination, the worse the self-rated health status of the elderly.

Aging attitude is the view and evaluation of the elderly on their own or others’ aging, which can be divided into positive and negative aspects. A large number of studies show that long-term negative aging attitude of individuals will increase the risk of negative physiological reactions (Levy et al., 2002). When elderly individuals take a negative attitude towards their aging, it will directly hinder their physical and mental health development and lead to maladjustment of individuals in the later life (Manzi et al., 2023). A positive aging attitude will help the elderly release stress (Fu Tingting et al., 2020) and promote health. Based on this, this paper puts forward research hypothesis 2:

Research hypothesis 2: Aging attitude will positively affect the self-rated health of the elderly. The more positive the aging attitude, the better the self-rated health status of the elderly.

The influence of perceived age discrimination on the self-rated health of the elderly is not single and direct, but involves multiple factors and multi-level complex processes. The theory of social exclusion reveals that the elderly are increasingly becoming lonely and helpless groups because of being excluded by the mainstream society, and the consequences of this exclusion will gradually affect the elderly’s views on themselves and others’ aging, and ultimately affect their physical and mental health; In addition, the mediating role of aging attitude in related factors affecting the health of the elderly has also been confirmed by related studies (Shi Jiaming and Liu Xiaoting, 2024; Yin Lanxin et al., 2023). Other studies have found that the elderly who hold negative age stereotypes and perceptions will lower their self-esteem by feeling old and feeling worse about their aging (Marquet M et al., 2019), thus damaging their health. Based on this, we assume that perceived age discrimination will affect the self-rated health by affecting the aging attitude of patients. Based on this, this paper puts forward research hypothesis 3:

Research hypothesis 3: Aging attitude plays an intermediary role in the process of age discrimination perception affecting the self-rated health of the elderly.

The theory of community governance provides a theoretical framework for the development of elderly-friendly communities. In-situ pension can ensure that the elderly live in a familiar environment and have a familiar interpersonal network. In-situ pension can bring the elderly a sense of security and self-identity, which will have an impact on their aging attitude. A rich community welfare can improve the happiness of the elderly and make them feel cared for and cared for (Lu Xuanru et al., 2022; Berke et al., 2007) is more likely to form a positive aging attitude, so this paper assumes that community welfare can buffer or offset the negative impact of age discrimination perception on aging attitude to some extent. Therefore, this study proposes hypothesis 4:

Research hypothesis 4: Community welfare plays a moderating role in the path of age discrimination perception affecting the aging attitude of the elderly.

## Data and methods

### Data Source

The research data of this paper is from the social follow-up survey of the elderly in China in 2020. The sample in this paper comes from the national sampling data, so the conclusion has certain typical significance.

### variables

#### 1. Explained variable: self-rated health

Measured by the question set in CLASS 2020, "What do you think of your current physical health? Is it very healthy, relatively healthy, average, relatively unhealthy, or very unhealthy? " As a basis for the self-rated health of the elderly, its options include: very healthy, relatively healthy, average, relatively unhealthy and very unhealthy, as a criterion for measuring the self-rated health of the elderly.

#### 2. Explanatory variable: perception of age discrimination

Latent variable constructed from four items (Cronbach’s α = 0.754):

“Modern society is hard to adapt”

“More and more social views are hard to accept”

“More and more new social policies are hard to accept”

“Current social changes are increasingly unfavorable to the elderly”

(1=Completely inconsistent, 5=Completely consistent).

#### 3. Mediator: Aging attitude

Summed score of items assessing views on aging (e.g., “Getting old is a process of constant loss”, reverse-scored; “Wisdom grows with age”). Higher scores indicate more positive attitudes.

#### 4. Moderator: Community welfare satisfaction

Average score of satisfaction with: road conditions, fitness/activity places, public security, environmental sanitation, atmosphere of respecting the elderly, staff capacity of neighborhood committees, and barrier-free facilities (1=Very dissatisfied, 5=Very satisfied). Higher scores indicate higher satisfaction (better community welfare).

#### 5. Control variables

The control variables in this study include personal physiological characteristics and social characteristics of the respondents. Personal physiological characteristics include age, gender, etc. Social characteristic variables include marital status, nationality, education level, political party, registered permanent residence, work situation, monthly average expenditure, etc.

### Model setting

In this study, the structural equation model was used to explore the influence of age discrimination perception and aging attitude on the self-rated health of the elderly. Structural equation model includes two parts: structural model and measurement model, in which the structural model represents the relationship between the variables studied, and the measurement model represents the relationship between latent variables and endogenous explicit variables. The latent variable of this study is age discrimination perception, and the model is basically set as follows:

The structural model in the structural equation model is:

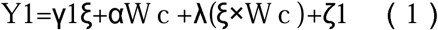

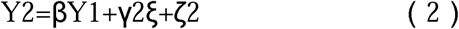

In formula (1), Y1 represents the aging attitude of the endogenous explicit variable, ξ refers to the perception of age discrimination of the exogenous latent variable, and W c represents the total score of community welfare after centralization, representing the deviation of the welfare level of the community where the individual is located from the average level of the sample. ξ×W c: it represents the interaction between age discrimination perception (ξ) and centralized community welfare (W c). α represents the main effect coefficient of community welfare (W c) on aging attitude (Y1). λ represents the core regulation effect coefficient. It is used to test whether community welfare (W c) regulates the influence intensity of age discrimination perception (ξ) on aging attitude (Y). If λ < 0 is significant, it shows that higher community welfare can weaken (buffer) the negative influence of age discrimination perception on aging attitude; If λ > 0 and significant, it indicates amplification effect. ζ1 represents the residual of structural model, which refers to the unexplained variance of aging attitude, and must satisfy cov(ξ,ζ)=0.

In Formula (2), Y1 represents the aging attitude of the endogenous explicit variable, Y2 represents the self-rated health of the endogenous explicit variable, and ζ 2 represents the residual of the structural model, which refers to the unexplained variance of the self-rated health, and must satisfy cov(ξ,ζ)=0.

The measurement model in the structural equation model is:

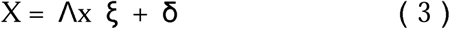

Equation (3) represents the measurement model of age discrimination perception of exogenous latent variables. Vector x represents the explicit variable index of exogenous latent variables. λ x represents the correlation (factor load) between the explicit variable and the latent variable ξ, and δ is the measurement error.

### Model construction

In this study, the explained variables, latent variables and series of control variables are included in the model. Based on the existing theoretical analysis, the aging attitude is set as an intermediate variable, and an intermediate model is constructed. The maximum likelihood estimation method (ML) is used for model fitting and result evaluation. Considering that social policies make it difficult for the elderly to accept and social changes are not good for the elderly, that is, when the elderly perceive that modern social policies are unacceptable, their adaptability to social changes will also decrease accordingly. In order to fit the facts better, the residual correlation line between "social policies are unacceptable" and "social changes are not good for the elderly" is added to the model.

Figure 2 reports the initial model results based on standardized coefficients. All the measured parameters of the initial model have passed the significance test of 0.1%, and the fitting indexes of the initial model are RMSEA=0.024, SRMR=0.010, CFI=0.996, and TLI=0.991, which all meet the fitting standards.

**Fig. 2.**
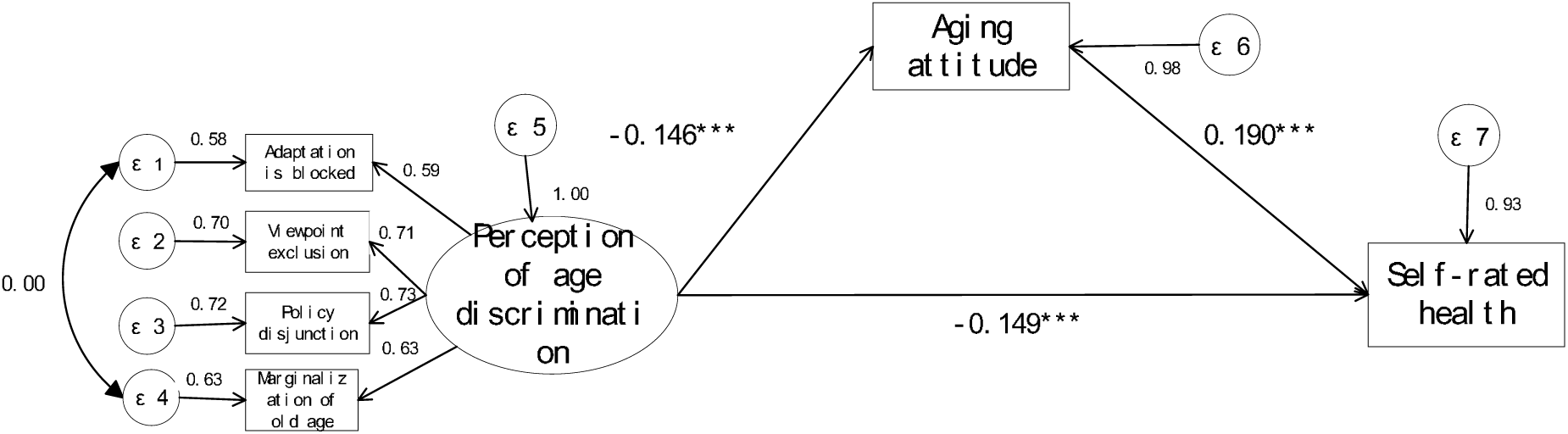
SEM model of the influence of age discrimination perception and aging attitude on self-rated health of the elderly. Note: standardization coefficient, * * p < 0.05, ** p < 0.01, *** p < 0.001.

## Results

### Sample Characteristics

Table 1 reports the descriptive statistical analysis results of the survey samples in this study. The sample age is between 60 and 98 years old, with an average age of 71.57 years old. The self-rated health status of all samples is concentrated in the range of "average" and "relatively healthy", accounting for 79.52%. In terms of gender distribution, there is little difference between male and female samples. The education level is concentrated in junior high school and below. 75.50% of the samples have spouses, 51.07% are non-agricultural registered permanent residence, and 3.36% are party member. The average monthly expenditure of all samples in the past 12 months is 2940.16 yuan. Of all the samples, 35.02% think that the current social changes are too fast to adapt, 35.11% think that the current views are increasingly unacceptable, 35.51% think that more and more new social policies are unacceptable, and 31.54% think that the current social changes are increasingly unfavorable to the elderly.

**Table 1.**
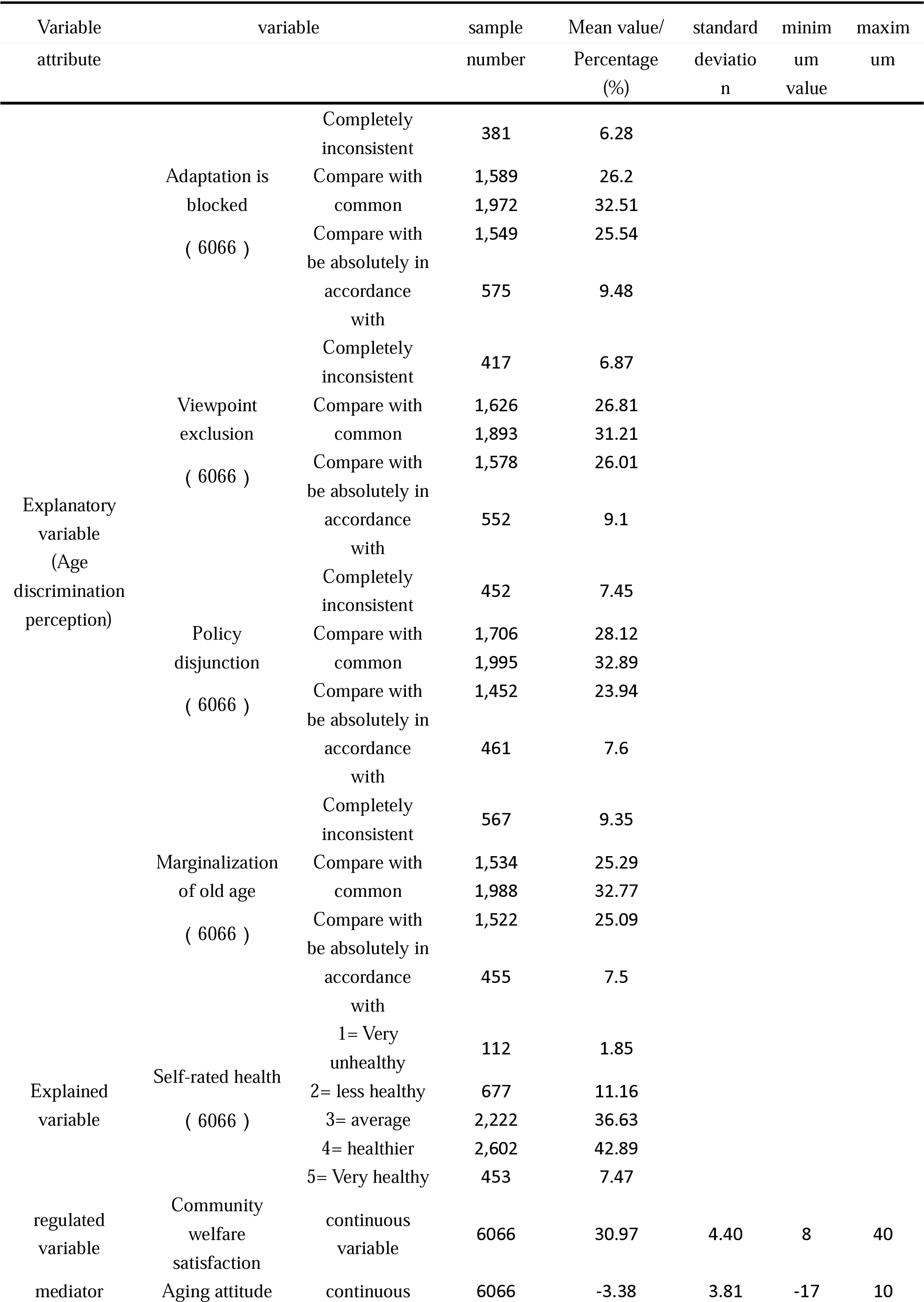
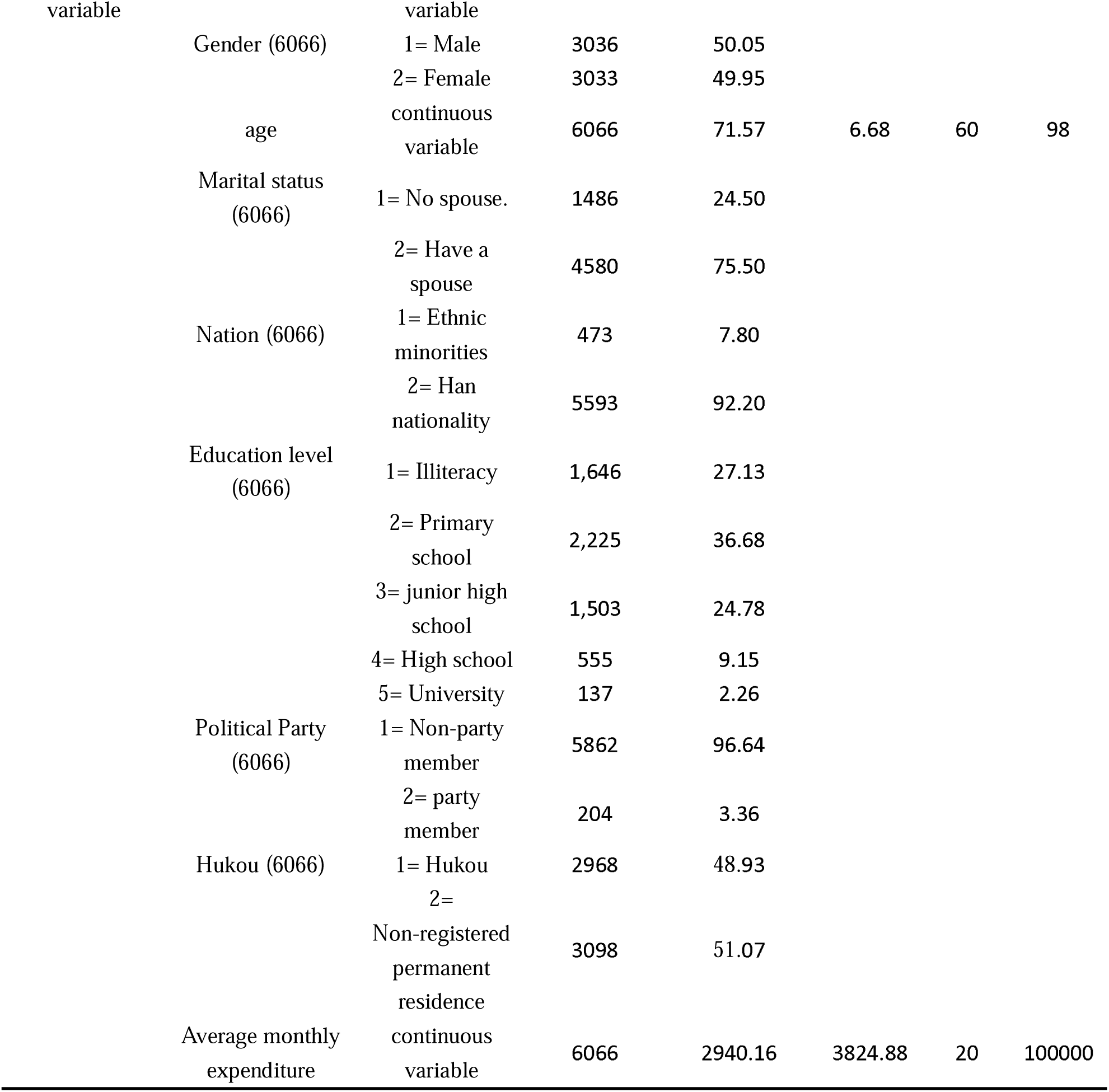
Descriptive Statistical Analysis of Samples.

### Main Effects

Table 2 reports in detail the estimation results of structural equation model that age discrimination perception and aging attitude affect the self-rated health of the elderly, which is divided into three parts: a. structural model, b. measurement model and c. fitting index.

**Table 2.**
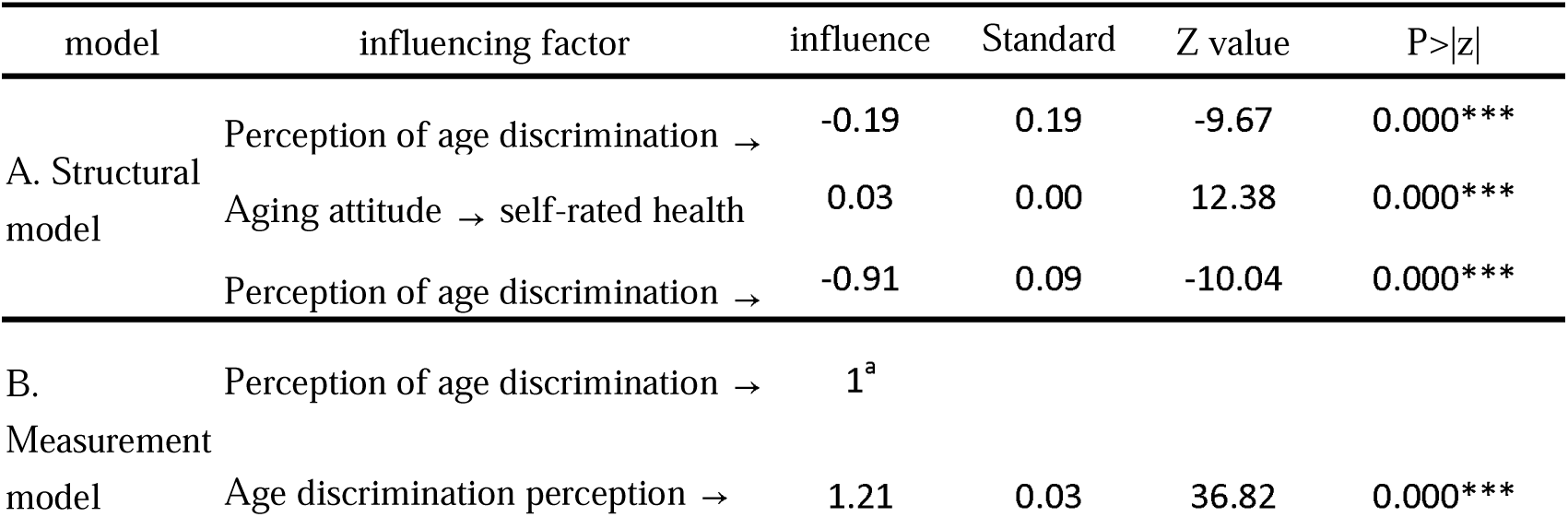
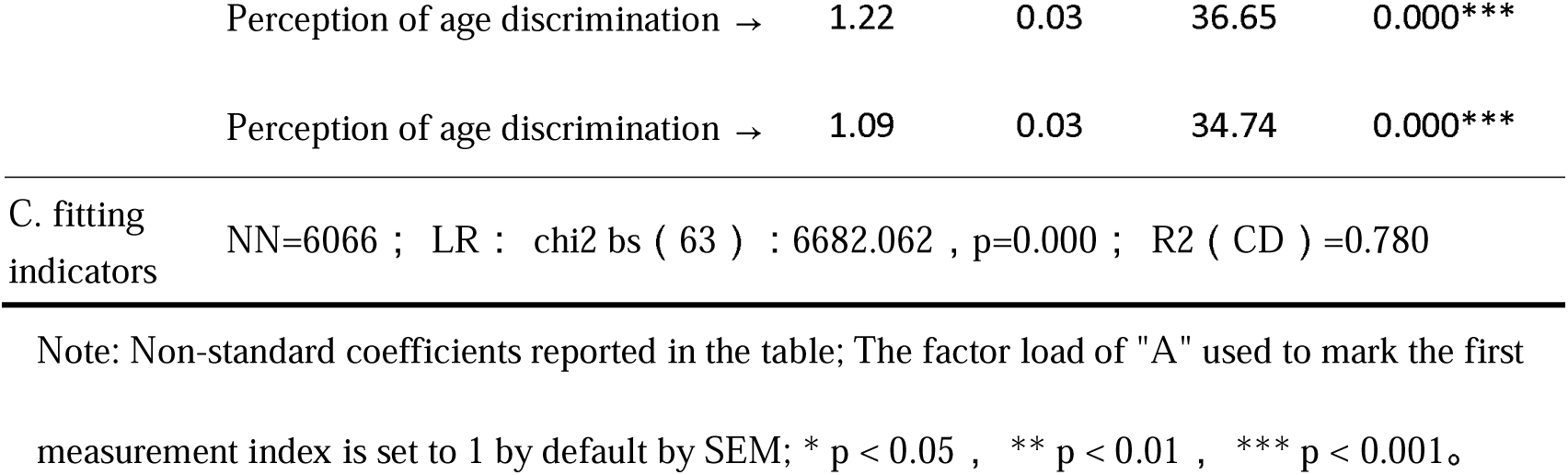
Structural Equation Model of the Influence of Age Discrimination Perception on Self-rated Health of the Elderly.

In a. structural model, the effects of age discrimination perception and aging attitude on the self-rated health of the elderly all passed the significance test and were significant at the level of 0.1%. Among them, the influence coefficient of age discrimination perception on self-rated health is -0.19, indicating that the stronger the age discrimination perception of the elderly, the worse their self-rated health status; The coefficient of influence of aging attitude on self-rated health is 0.03, which shows that the more positive the aging attitude of the elderly (their views and attitudes towards the aging phenomenon of themselves and others tend to be positive), the better their self-rated health. The influence of age discrimination perception on aging attitude also passed the significance test, and it was significant at the level of 0.1%, with the influence coefficient of -0.91, indicating that the more obvious the perception of age discrimination, the more negative the aging attitude of the elderly (the views and attitudes towards themselves and others aging tend to be negative).

C. In the fitting index, the RMSEA of the model RMSEA=0.031, which is less than the acceptable upper limit of the model fitting degree of 0.08; CFI=0.964, TLI=0.944, and the model fits well. R2(CD) of the whole model is 0.780, which means that the explanatory power of the whole equation is 78%, that is, 78.5% of the total variance of all endogenous variables (including potential variables and observed variables) in the model is jointly explained by all predictive variables (exogenous variables and endogenous variables) in the model. It shows that this model can be accepted to explain the influence of age discrimination perception on the self-rated health of the elderly.

Figure 3 shows the main effects of age discrimination perception and aging attitude on self-rated health. Specifically, the perception of age discrimination has a negative impact on the self-rated health and aging attitude of the elderly, and it is significant at the level of 0.1%. It shows that the stronger the perception of age discrimination among the elderly, the worse their self-rated health status and the more negative their attitude towards aging (their views and attitudes towards themselves and others tend to be negative). Aging attitude has a positive effect on the self-rated health of the elderly, and it is significant at the level of 0.1%. It shows that the more positive the aging attitude of the elderly (the view and attitude towards the aging phenomenon of themselves and others tend to be positive), the better their self-rated health status.

**Fig. 3.**
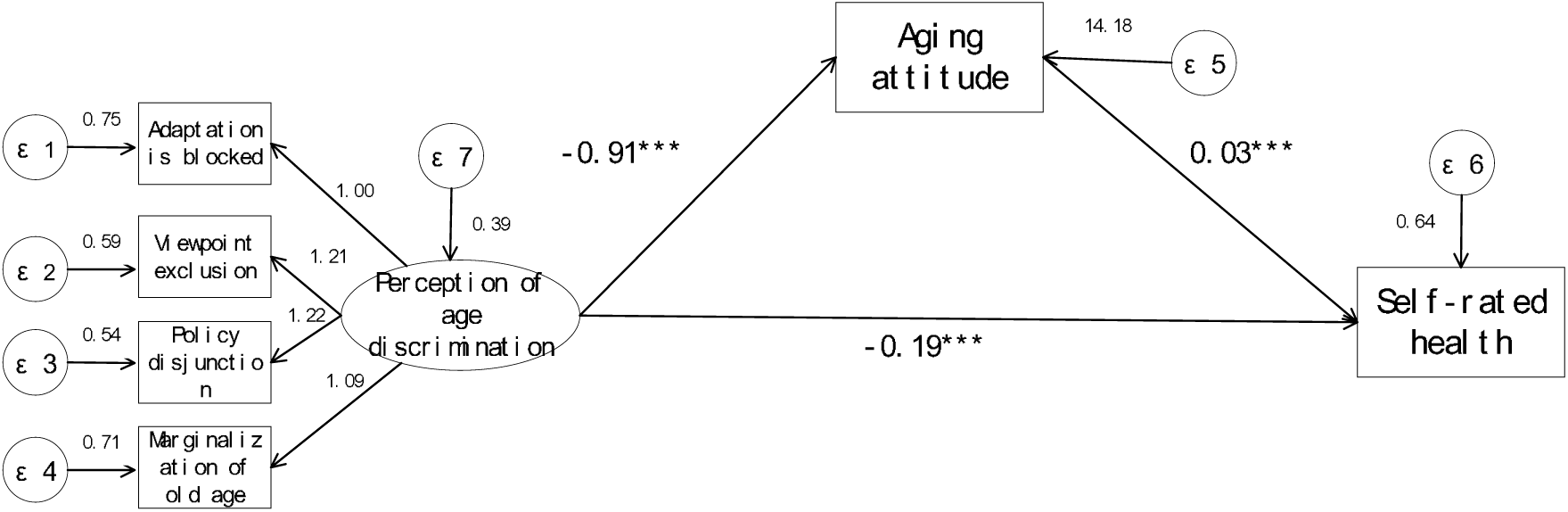
Main effect analysis of age discrimination perception and aging attitude on self-rated health.

### Mediation Effect

In addition to the influence of age discrimination perception and aging attitude on the self-rated health of the elderly, this study also pays attention to the intermediary role of aging attitude in the influence of age discrimination on self-rated health. This research model contains two paths that age discrimination perception affects self-rated health, namely L direct path: age discrimination perception→ self-rated health, L indirect path: information perception → aging attitude → self-rated health.

Table 3 reports the direct effect, indirect effect and total effect among the variables in the model. As shown in Table 4-7, the direct effect and total effect of all paths in the model are significant at the level of 0.1%, indicating that the perception of age discrimination does have a significant negative impact on the self-rated health of the elderly; The indirect effect value of the intermediary effect of aging attitude is -0.032, which is significant at the level of 0.1%. However, testing the significance of indirect effect coefficient is not enough to prove the existence of indirect effect, so this study continues to test the significance of coefficient product of indirect path. Sobel test and Bootstrap test are commonly used to test the significance of coefficient product, and their standards are respectively: if the z value of Sobel test reaches the significance level of 5% and the confidence interval of Bootstrap test does not include 0, the indirect effect is regarded as significant (Fang Jie et al., 2010).

**Table 3.**
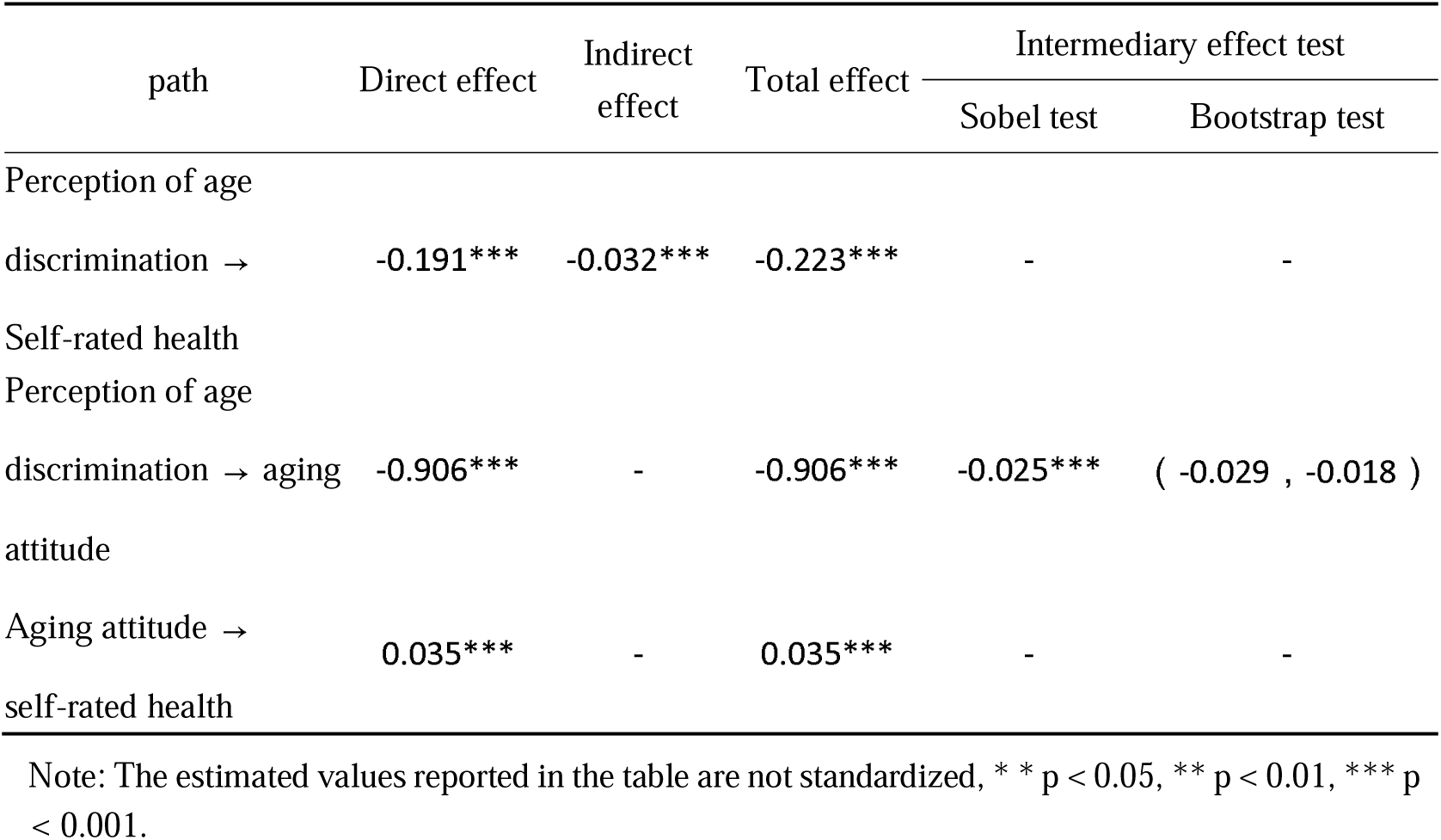
Model Effect Decomposition and Intermediary Test.

**Table 4.**
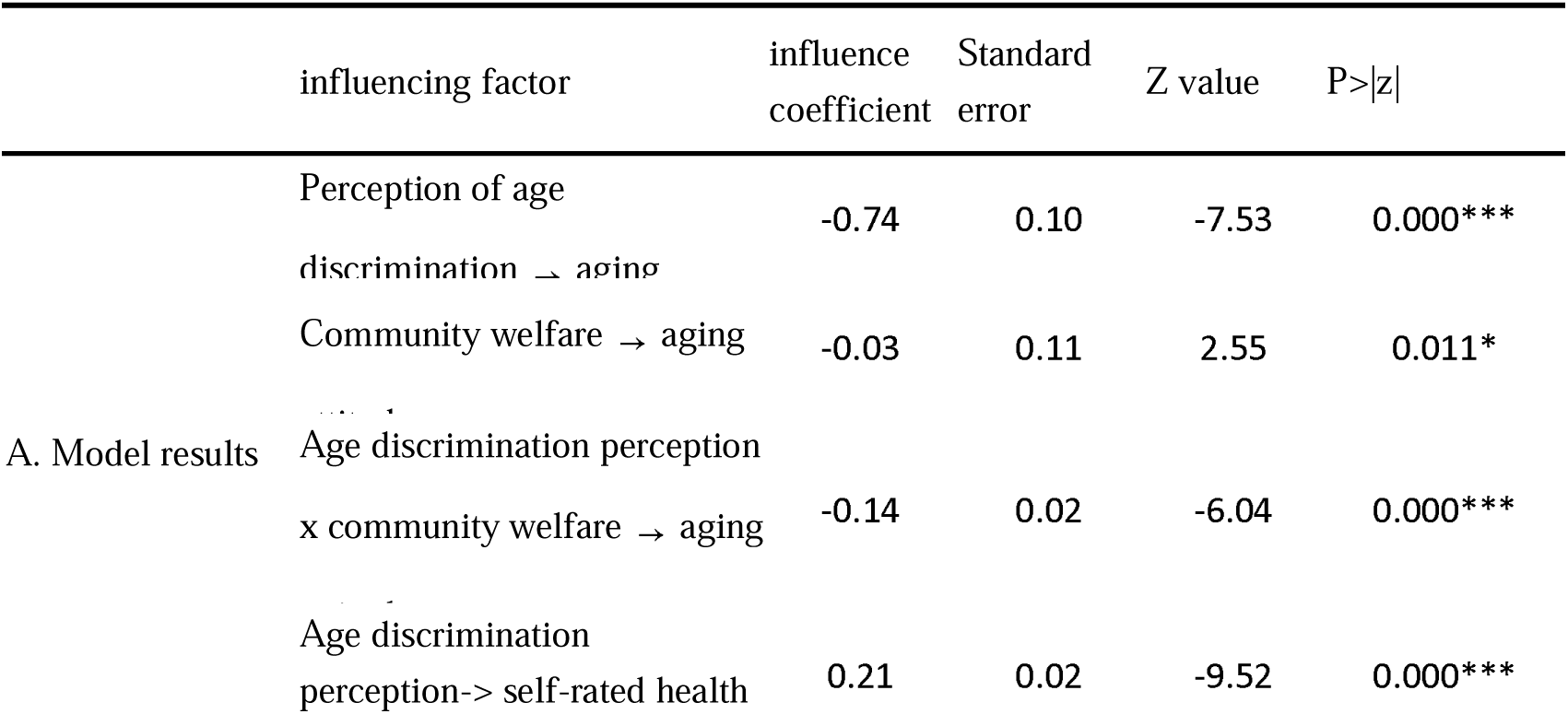
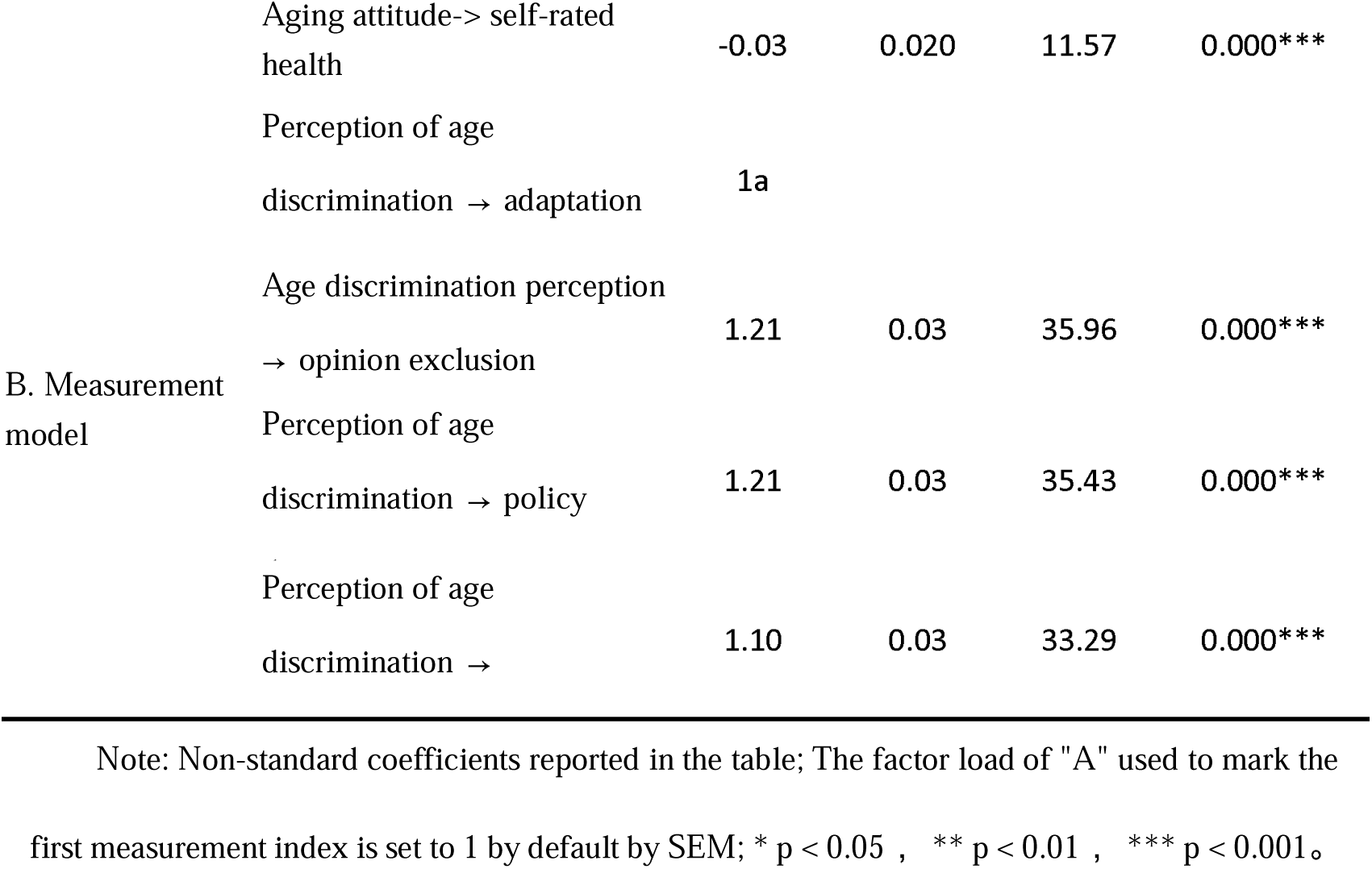
Test of the moderating effect of community welfare on the perception of age discrimination affecting the aging attitude of the elderly.

Specifically, in Figure 4, in the two paths of aging attitude, the Sobel test result of coefficient product is significant at the level of 0.1%, and the confidence interval of Bootstrap test is (-0.029, -0.018), and 0 is not within the confidence interval. According to the criterion of mediation effect, this shows that there is a significant partial mediation effect in aging attitude. That is to say, in the indirect path of "age discrimination perception-aging attitude-self-rated health", the influence of age discrimination perception on self-rated health occurs partly through aging attitude, and there are also other unobserved intermediary paths. Combined with the principal effect analysis of the model, we can find that the perception of age discrimination can not only directly negatively affect the self-rated health of the elderly, but also indirectly reduce their self-rated health by affecting their aging attitude. Figure 7 visually shows the mediating effect of age discrimination perception on self-rated health.

**Fig. 4.**
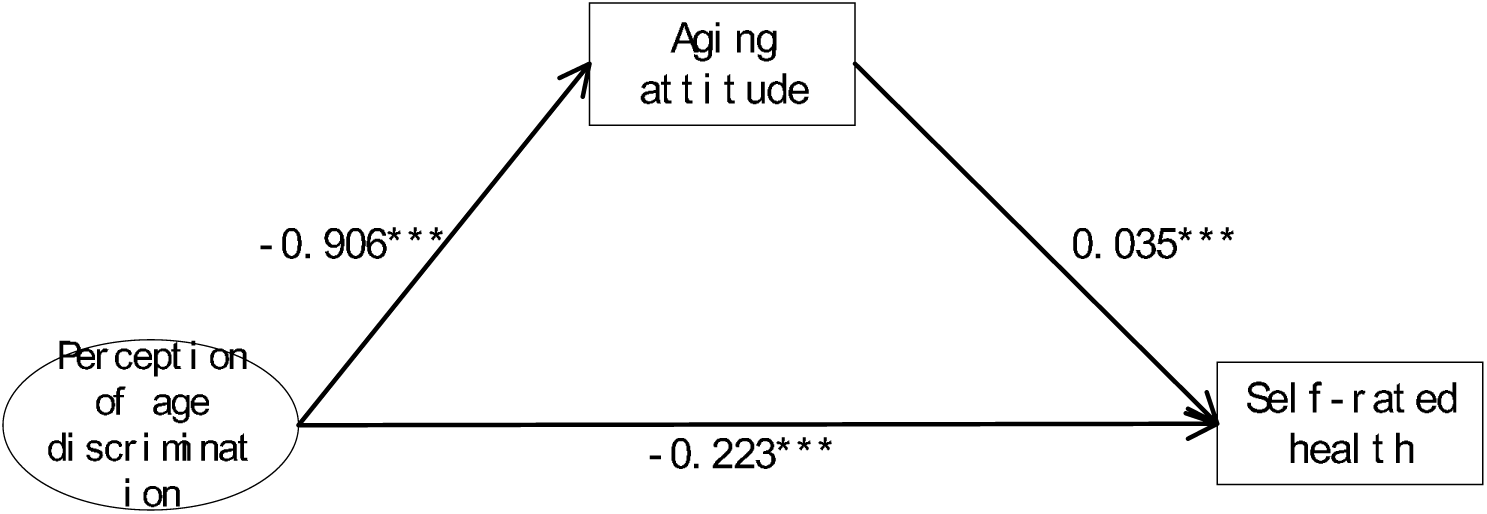
Analysis of the Mediating Effect of Age Discrimination Perception on Self-rated Health.

### Moderation Effect

SEM can well deal with the model containing regulatory effects, especially the interaction effects of latent variables. When at least one model is a latent variable, the analysis method of latent variable regulation effect should be used. Among them, when the independent variables are latent variables and the regulatory variables are explicit variables, the product index method under the framework of structural equation model (SEM) should be used to test the regulatory effect in Stata.

Table 4 reports the moderating effect of age discrimination perception on the aging attitude of the elderly in community welfare. It can be seen from the results that the influence coefficient of age discrimination perception on aging attitude is -0.63, and it is significant at the level of 0.1%. This shows that the more age discrimination the elderly perceive, the more negative their aging attitude will be. Every time the perception of age discrimination increases by 1 standard deviation, the aging attitude score decreases by 0.63 standard deviation on average. The influence coefficient of the interaction between age discrimination perception and community welfare on the aging attitude of the elderly is -0.13, and it is significant at the level of 0.1%. This shows that community welfare has indeed played a positive role in regulating the path of age discrimination perception affecting the aging attitude of the elderly. That is, when the level of community welfare is high, the negative impact on aging attitude will be reduced by 0.13 unit for every unit of age discrimination perception. It is also worth noting that the influence coefficient of community welfare on aging attitude is -0.03, and it is significant at the level of 1%. This shows that although community welfare plays a positive regulatory role in the path of age discrimination perception affecting the aging attitude of the elderly, it has a negative and significant impact on the aging attitude itself, and the specific reasons will be discussed later.

Figure 5 shows the moderating effect of community welfare in the first half of intermediary effect. Specifically, compared with the results shown in Figure 4-3, although community welfare itself has a negative impact on aging attitude, the negative impact of age discrimination perception on aging attitude has indeed declined after community welfare has played a regulatory role, and finally the negative impact of age discrimination perception on self-rated health has also declined, and the result is significant at the level of 0.1%. This shows that community welfare can significantly negatively regulate the negative impact of age discrimination perception on the aging attitude of the elderly, that is, the higher the level of community welfare, the smaller the negative impact of age discrimination on the aging attitude of the elderly, and ultimately the smaller the negative impact on the self-rated health of the elderly.

**Fig. 5.**
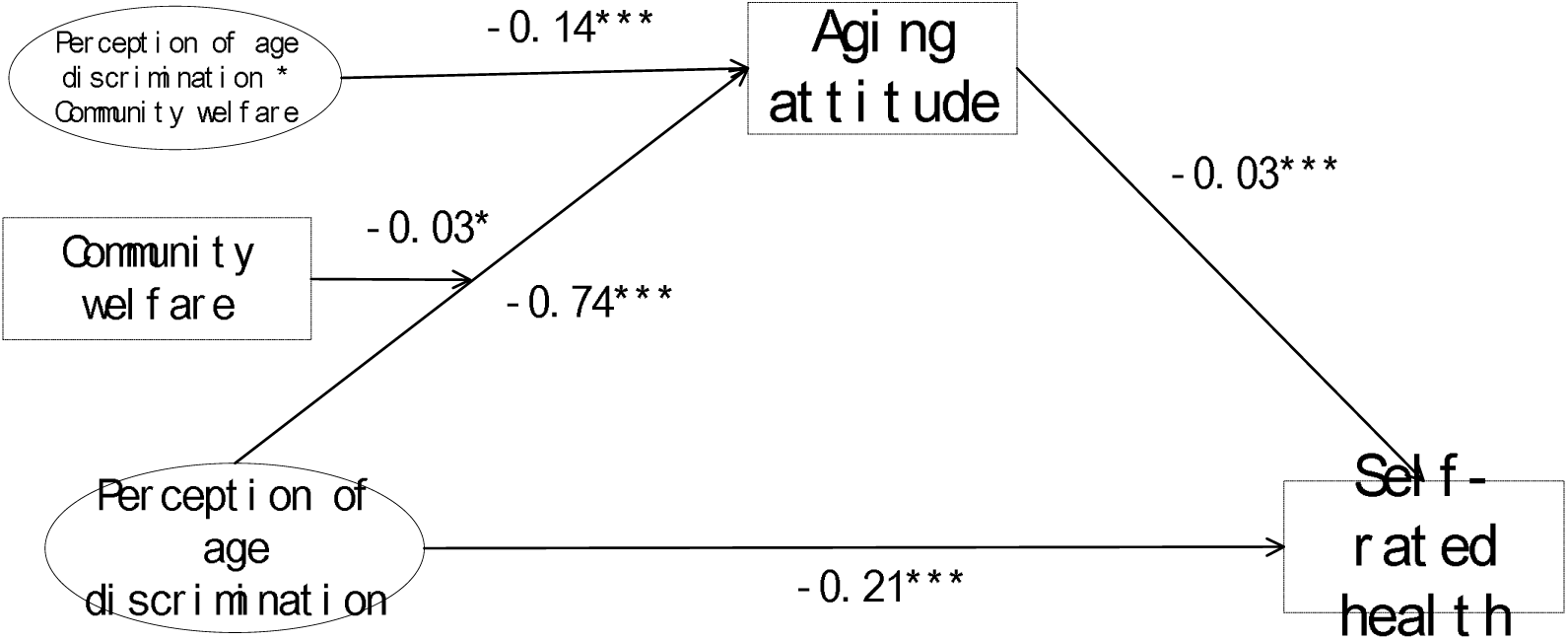
The moderating effect of age discrimination perception on the aging attitude of the elderly in community welfare.

### Robustness Checks

In this study, two estimation methods are used to test the robustness of the above model results: (1) Robust standard error (ML+Robust) based on maximum likelihood estimation and heteroscedasticity; (2) Bootstrap method (1000 Bootstrap samples) was adopted.

Table 5 reports the results of the robustness test. From the results, it can be seen that there is not much difference between the two methods in the influence coefficient of each path. The significance level of the path coefficient of age discrimination perception affecting self-rated health is still 0.1%, which shows that the negative influence of age discrimination perception on self-rated health still exists significantly. The significance level of the path coefficient of aging attitude affecting self-rated health is still 0.1%, which shows that the positive influence of aging attitude on self-rated health still exists significantly. It can be seen that the structural equation model constructed in this study reports the same results under different estimation methods and has certain robustness.

**Table 5.**
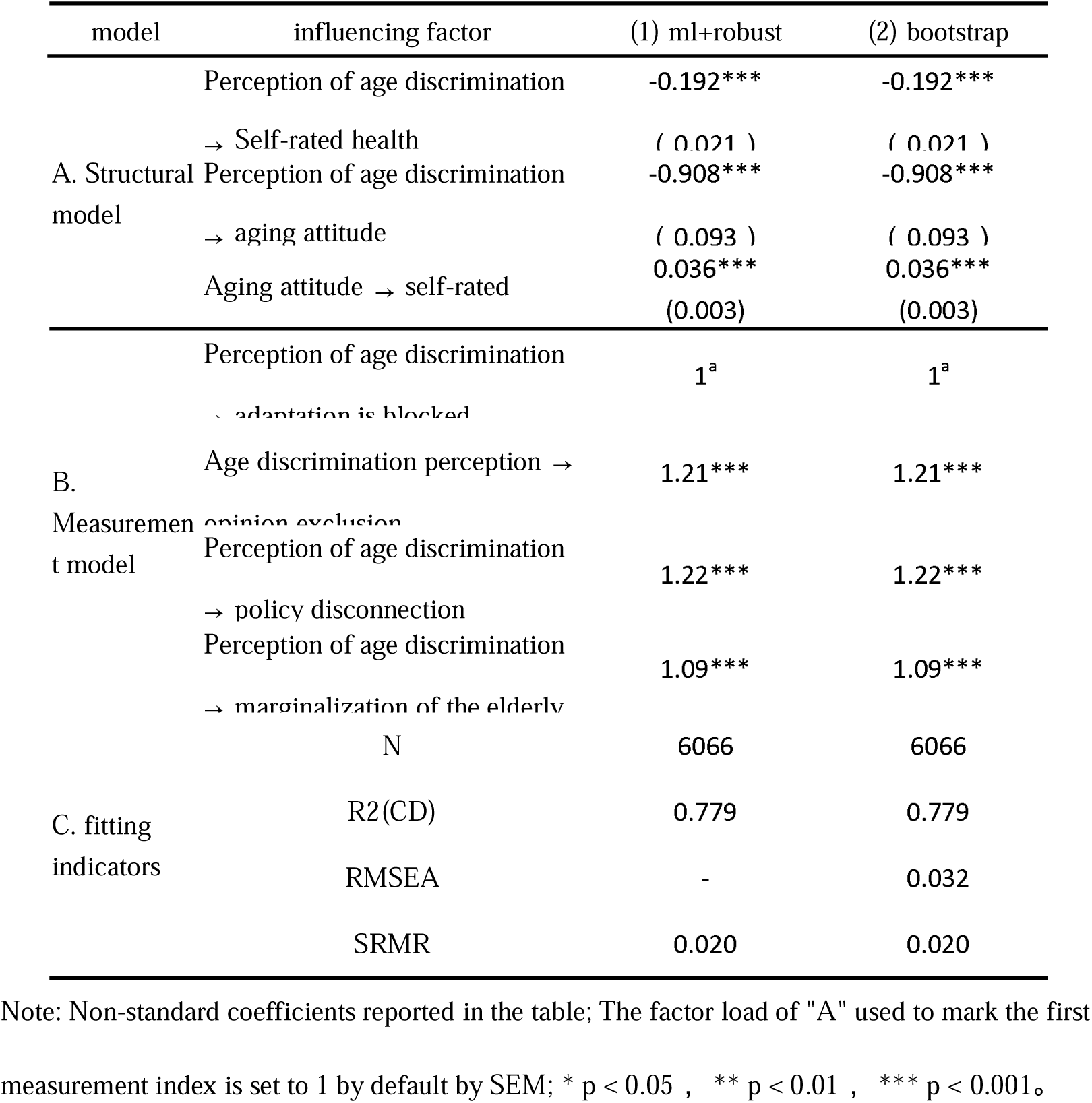
SEM Robustness Test on the Influence of Age Discrimination Perception on Self-rated Health of the Elderly.

## Discussion

This study investigated the mechanism linking age discrimination perception to self-rated health in Chinese older adults, focusing on the mediating role of aging attitude and the moderating role of community welfare. Key findings are:

H1 Confirmed: Age discrimination perception directly and negatively impacts self-rated health, aligning with global findings (Levy et al., 2023; Jackson et al., 2019) and social exclusion theory, where discrimination marginalizes seniors, harming well-being.

H2 Confirmed: A positive aging attitude significantly improves self-rated health. Negative self-perceptions of aging act as stressors, hindering health, while positive views promote resilience and better health management (Levy et al., 2002; Manzi et al., 2023; Fu Tingting et al., 2020).

H3 Confirmed: Aging attitude acts as a significant partial mediator. Age discrimination erodes positive views on aging, which in turn worsens health. This mechanistic insight is novel for this relationship.

H4 Partially Confirmed: Community welfare buffers the negative impact of age discrimination on aging attitude. This supports the potential of elderly-friendly communities (Lv xuanru et al., 2022; Berke et al., 2007). However, the unexpected small negative direct effect of community welfare on aging attitude warrants investigation. It might reflect feelings of dependency fostered by welfare provision ("being cared for implies weakness").

## Implications

Policy: Strengthen anti-age discrimination laws and actively promote positive aging narratives. Invest in comprehensive community welfare (social activities, accessible environments, respectful care) to buffer discrimination effects.

Practice: Develop community programs fostering positive aging attitudes (discussion groups, intergenerational activities). Train community staff to deliver services that empower rather than induce dependency.

Research: Explore other mediators (e.g., social isolation, self-efficacy) and moderators. Investigate the counterintuitive direct effect of community welfare. Use longitudinal designs.

## Strengths and Limitations

Strengths: Large nationally representative sample; SEM modeling of latent variables; robust moderated mediation analysis.

Limitations: Cross-sectional data limits causal inference. Self-reported measures susceptible to bias. Unexplored mediators/moderators. Generalizability to rural or institutionalized elderly may be limited.

## Conclusion

This study demonstrates that age discrimination perception negatively impacts older adults’ self-rated health both directly and indirectly through its detrimental effect on aging attitudes. Community welfare emerges as a crucial protective factor, mitigating the negative impact of discrimination on aging attitudes. These findings underscore the urgent need to combat age discrimination through policy and public awareness campaigns, actively promote positive aging narratives, and prioritize the development of empowering community welfare programs designed to foster autonomy and well-being, not dependency, among the elderly population in China.

## Data Availability

The data underlying the results presented in the study are available from http://class.ruc.edu.cn/xmjs/xmgk.htm. The China elderly social survey (CLASS) is a nationwide and continuous large-scale social survey project. By regularly and systematically collecting the social and economic background data of the elderly population in China, we can grasp the problems and challenges faced by the elderly in the aging process, evaluate the actual effects of various social policies and measures in improving the quality of life of the elderly, and provide important theoretical and factual basis for solving the aging problem in China.

http://class.ruc.edu.cn/index.htm

